# Antibody escape, the risk of serotype formation, and rapid immune waning: modeling the implications of SARS-CoV-2 immune evasion

**DOI:** 10.1101/2023.01.25.23285031

**Authors:** Catherine Albright, Debra Van Egeren, Aditya Thakur, Arijit Chakravarty, Laura F. White, Madison Stoddard

## Abstract

As the COVID-19 pandemic progresses, widespread community transmission of SARS-CoV-2 has ushered in a volatile era of viral immune evasion rather than the much-heralded stability of “endemicity” or “herd immunity.” At this point, an array of viral variants has rendered essentially all monoclonal antibody therapeutics obsolete and strongly undermined the impact of vaccinal immunity on SARS-CoV-2 transmission. In this work, we demonstrate that antigenic drift resulting in evasion of pre-existing immunity is highly evolutionarily favored and likely to cause waves of short-term transmission. In the long-term, invading variants that induce weak cross-immunity against pre-existing strains may co-circulate with those pre-existing strains. This would result in the formation of serotypes that increase disease burden, complicate SARS-CoV-2 control and raise the potential for increases in viral virulence. Less durable immunity does not drive positive selection as a trait, but such strains may transmit at high levels if they establish. Overall, our results draw attention to the importance of inter-strain cross-immunity as a driver of transmission trends and the importance of early immune evasion data to predict the trajectory of the pandemic.

## Introduction

Some early commentators bullishly predicted the end of the COVID-19 pandemic [1–4], with the build-up of vaccine and natural immunity eventually curtailing SARS-CoV-2 transmission. However, the pandemic is now entering its fourth year despite a vast burden of prior infection, over 13 billion vaccination doses globally [5] and high prevalence of anti-SARS-CoV-2 antibodies [6,7]. Consistent with early-pandemic warnings [8–13], the pace of immune evasion has proven rapid [14,15], and transmission has continued robustly in the post-vaccine era [16]. Warnings about insufficient vaccine acceptance [17], rapid waning of vaccine and post-infection immune protection [18,19], and antibody evasion [8–10,20] have all materialized at this point [21–23], leading to the high levels of SARS-CoV-2 transmission.

The ability of SARS-CoV-2 to evade immunity through mutations that degrade antibody binding has been a major driver of high and variable viral transmission. Indeed, the post-omicron era of the pandemic has been marked by successive waves driven by immune-evading subvariants, including BA.1, BA.5, XBB, and XBB.1.5 [14,22,24–26]. These immune-evading strains acquire an evolutionary advantage in the context of widespread immunity through mutations that degrade the binding of antibodies induced by infection with prior strains or by vaccines (“antigenic drift”)[27]. As neutralizing antibodies mediate sterilizing immunity to SARS-CoV-2–that is, they block infection upon exposure—evasion of these antibodies promotes reinfection [28]. This advantage has allowed these immune-evading strains to achieve dominance, drive spikes in transmission, and replace (succeed) pre-existing strains [20]. Between December 2021 (initial dissemination of the original BA.1 omicron variant) and December 2022, several strain succession events were documented resulting in an approximately 35-fold loss in neutralizing titer [29].

Understanding the potential of emerging strains to drive waves of infection, persist in circulation, and co-circulate with pre-existing strains is vital for understanding and reacting to this volatile phase of the pandemic. Immune evasion has implications for short-term and long-term transmission levels [30], possible changes in disease severity (manuscript in preparation), and the efficacy of vaccines and therapeutics, especially monoclonal antibodies [22,31]. Anticipating the behavior of viral variants has tremendous practical significance for designing nonpharmaceutical and biomedical interventions.

In this study, we use an epidemiological modeling framework to build a quantitative understanding of the role of immune evasion in inter-strain competition and selection dynamics under endemic conditions. To this end, we developed a two-strain Susceptible-Infectious-Recovered-Susceptible (SIRS) model accounting for variable cross-immunity between a pre-existing and an invading strain. This paper explores viral evolutionary strategies by simulating a few relevant immunological scenarios: antigenic drift, which we surmise may be symmetric or unilateral, and induction of less durable immunity.

Antigenic drift results in reduced cross-immunity (immunity induced by one strain against another) compared to homologous immunity (immunity induced by a strain against itself) [32]. If the impact of antigenic drift is symmetric, the invading strain’s cross immunity against the original strain will equal the original strain’s cross immunity against the invading strain. The plausibility of this scenario is supported by the tolerance of SARS-CoV-2’s spike for a wide variety of mutations [8,33,34]. However, omicron BA.1 appeared to benefit from essentially unilateral antigenic drift: while BA.1 strongly evaded pre-existing immunity to delta, the delta was impeded by immunity induced by BA.1 [35]. The final scenario regards the possibility of viral strains with reduced durability of immunological protection from reinfection. Possibly exemplifying this scenario is omicron, which appears to exert weaker protection against homologous reinfection than delta (prior omicron reduces risk of omicron reinfection by 59.3%; prior delta infection reduces risk of delta reinfection by 92.3% [36].)

Determining the immunological properties likely to be selected for is crucial for predicting the trajectory of SARS-CoV-2 under widespread transmission. Although the rapid pace of SARS-CoV-2 evolution and the simultaneous emergence of immune-evading multiple variants paints a complex picture [37,38], this simplified analysis provides a basis for understanding the inter-strain competition and selection that underpin these dynamics. Identifying the characteristics of strains likely to be successful and drive significant waves of transmission is crucial to support early-warning systems. Co-circulation of viral serotypes – that is, viral strains sufficiently antigenically distinct to coexist [39] – is an emergent threat that requires greater understanding and may lead to dramatically worse outcomes in the long-term trajectory of the pandemic.

## Methods

To evaluate short-term and long-term transmission of novel strains of SARS-CoV-2, we built a two-strain SIRS (susceptible-infectious-recovered-susceptible) model. The model has two sets of parallel compartments, representing those infected with the original strain and the invading strain. The model considers disease transmission, waning immunity, and induction of immunity due to exposure to either strain or both strains. Figure 1 is a visual representation of the model compartments and transitions between compartments through infection, recovery, and immune waning.

**Figure 1.**
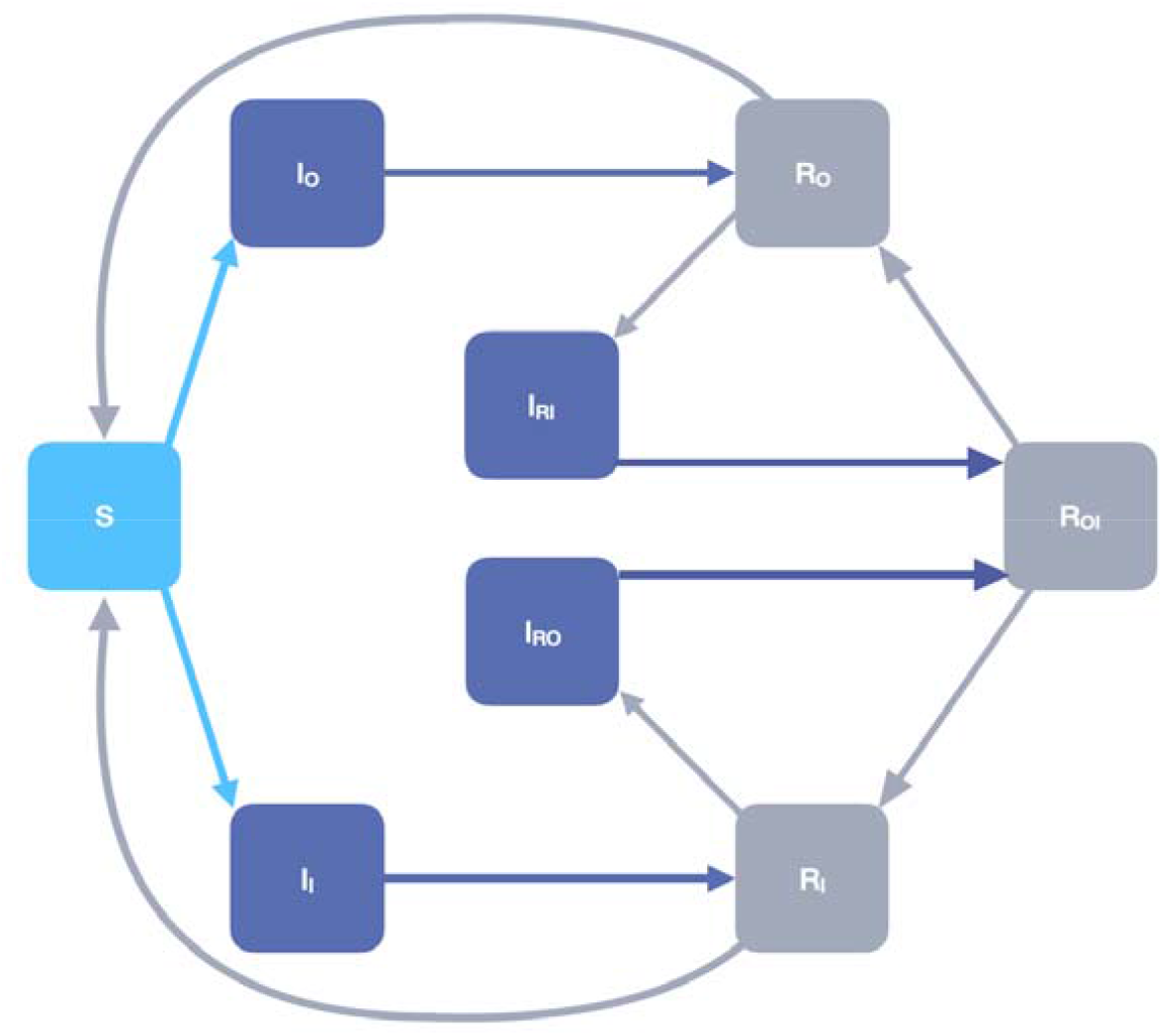
Schematic representing the two-strain SIRS model. Light blue represents fully susceptible individuals, dark blue represents infected individuals, and gray represents recovered individuals.

The model parameters that are fixed among all model iterations are listed in Table 1A; variable parameters and their ranges are described in Table 1B.

**Table 1A:** Model parameters fixed between scenarios.

| Symbol | Parameter | Value | Units |
| --- | --- | --- | --- |
| $\mu$ | Birth rate | $2.5 \times 10^{-5}$ [40] | 1/person/days |
| N | Population size | 330,000,000 [41] | Individuals |
| $\rho$ | Immune waning rate | 0.0018 [42] | 1/days |
| $\gamma$ | Recovery rate | 0.1 [43] | 1/days |
| $R_{0,o}$ | Intrinsic reproductive number, original strain | 8.2 [44,45] | Individuals |

The immune waning rate of 0.0018/day corresponds to a mean duration of protection from infection of 550 days.

**Table 1B:** Variable parameters and their values by scenario.

| Symbol | Parameter | Value, symmetric antigenic drift | Value, unilateral antigenic drift | Value, shorter duration of immunity | Units |
| --- | --- | --- | --- | --- | --- |
| $R_{0,i}$ | Intrinsic reproductive number, invading strain | 0 – 20 | 0 – 20 | 0 – 20 | individuals |
| $C_{OVI}$ | Cross-immunity of original strain against invading strain | 0 – 1 | 0 – 1 | 1 | n/a |
| $C_{IVO}$ | Cross-immunity of invading strain against original strain | 1 | 0 – 1 | 1 | n/a |
| $\tau$ | Waning factor | 1 | 1 | 1-10 | n/a |

The effective infectiousness for the original and invading strains, β_O_ and β_I_, respectively, are derived from the relationship between R_0,O_ and R_O,I_ and the recovery rate, γ. β_O_ and β_I_ are calculated as follows:

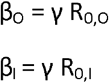

The effective infectiousness for reinfection, β_RO_, and β_RI_ can be derived from the relationship between the effective infectiousness and cross-immunity parameter, C:

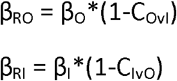

To represent symmetric antigenic drift, the cross-immunity of the original strain against the invading strain (C_OvI_) is set equal to the cross-immunity of the invading strain against the original strain (C_IvO_). In the shorter duration of immunity scenario, there is assumed to be complete cross-immunity between the two strains; as a result, no individuals are simultaneously immune to both strains (no individuals in the R_OI_ compartment).

The ordinary differential equations for the model are summarized below:

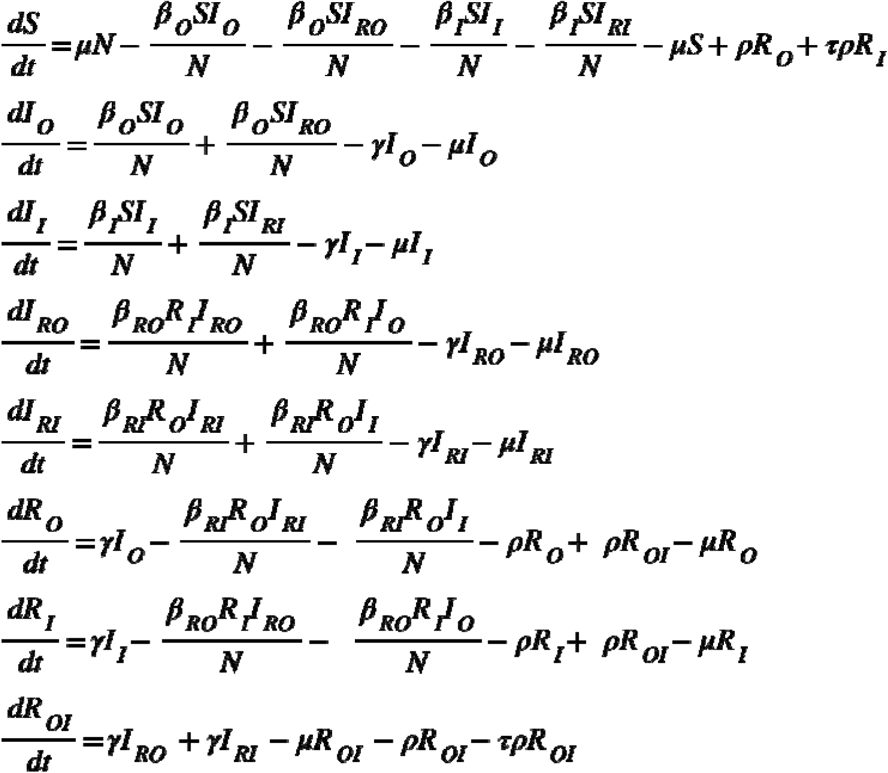

Where S is the susceptible population, I is the infectious population, and R is the recovered population. Subscripts O and I represent the original strain and the invading strain, respectively. Individuals in the R_OI_ compartment have immunity against both strains. I_rO_ represents individuals who are infected with the original strain while immune to the invading strain; I_rI_ represents individuals who are infected with the invading strain while immune to the original strain.

Initial conditions for the model are set such that the original strain is at an endemic steady-state, meaning the number of active infections is constant. The steady-state distribution of individuals in each compartment was determined by running the simulation for 10,000 days in the absence of the invading strain. Using this method, we found that the steady-state number of active original strain infections was 5.27 × 10, with 2.84 × 10 individuals recovered. To simulate invasion, we set the initial value of I_I_ – infections with the invading strain – to one.

We used the model to assess the invading strain’s ability to invade and drive a wave of transmission in the short-term as well as the impact of invading strains on long-term transmission trends. For the short term, we define an outbreak to succeed if the infected population ever increases above the initial value during the first 180 days. To determine long-term transmission trends, we ran the two-strain model for 10 years, after which point equilibrium is reached. Yearly infections are read out over the last year of the simulation.

## Results

### Mutual immune evasion favors successful invasion and co-circulation

In Figures 2 and 3, we explore epidemiological outcomes after the introduction of an invader strain with mutually reduced cross-immunity with respect to the original strain. We assumed immune evasion is perfectly symmetric: that is, the invader strain’s sensitivity to immunity induced by the original strain is equal to the original strain’s sensitivity to new immunity induced by the invader strain. Figure 2 demonstrates that if immune evasion is symmetric, invasion is successful under a wide range of transmissibility and immune evasion conditions – clearly, symmetric immune evasion is a successful strategy. Even a poorly transmissible strain relative to currently circulating variants may successfully invade if cross-immunity is low enough.

**Figure 2.**
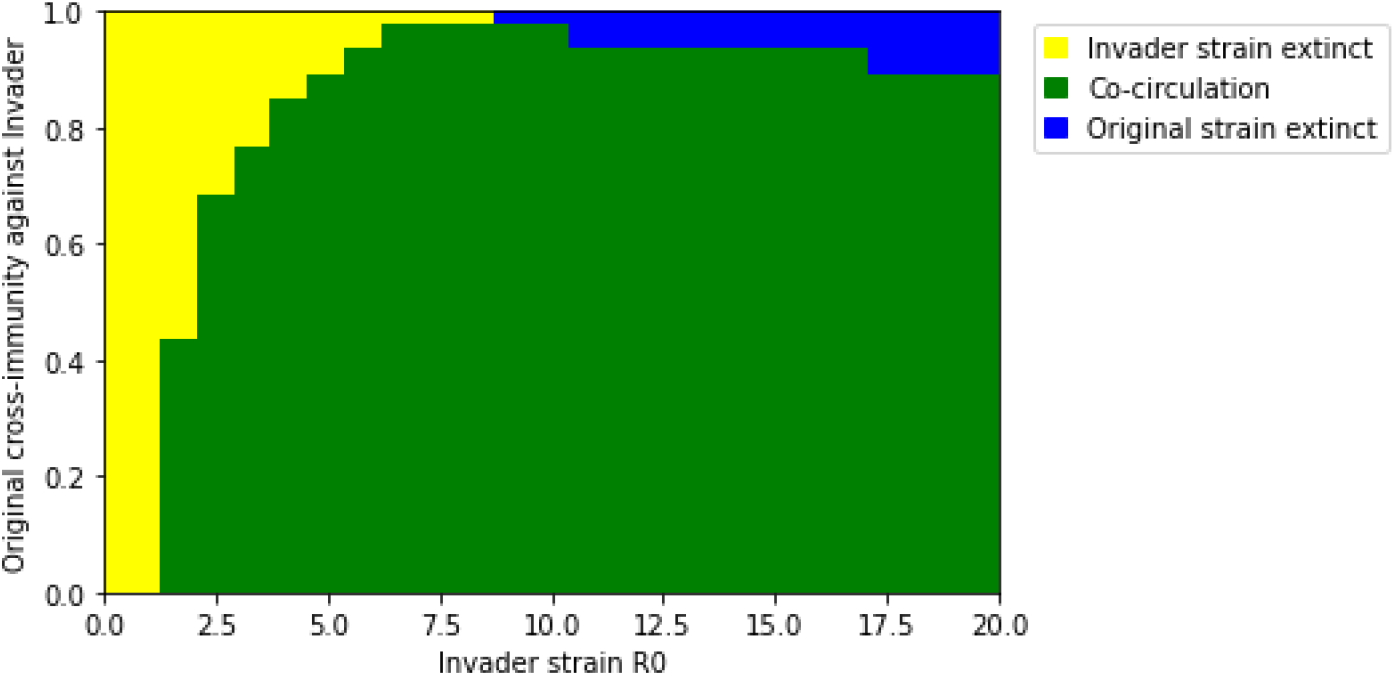
Invasion outcomes for invader strains with varying transmissibility (R_0_) and symmetric cross-immunity with the original strain. Outcomes are classified based on whether either strain becomes extinct. Invasion is successful unless the invader strain becomes extinct (yellow).

The success of the invader strain may not come at the expense of the original strain. Succession (extinction of the original strain) is expected only in scenarios where cross-immunity between the strains is high, and the invading strain is more transmissible than the original strain.

### Invasion increases transmission in the short-term, while co-circulation increases transmission in the long-term

Figure 3 explores short-term and long-term strain transmission of the original and invader strains after various invasion scenarios. As shown in Figure 3A, the impact of invasion on short-term transmission of the original strain may be modest, while extensive transmission of the invading strain may occur (Figure 3B). This results in a large increase in overall short-term transmission (a “wave” or a “spike”) for successful invasion scenarios compared to failed invasions, which reflect baseline transmission of the original strain (Figure 3C). Higher transmissibility of the invader strain and greater mutual immune evasion (lower cross-immunity) promote larger waves of transmission. In the long-term, symmetrically low cross-immunity resulting in co-circulation of strains results in the highest overall transmission (Figure 3D-F.) This is because as cross-immunity levels decrease, competition between the strains also decreases.

**Figure 3.**
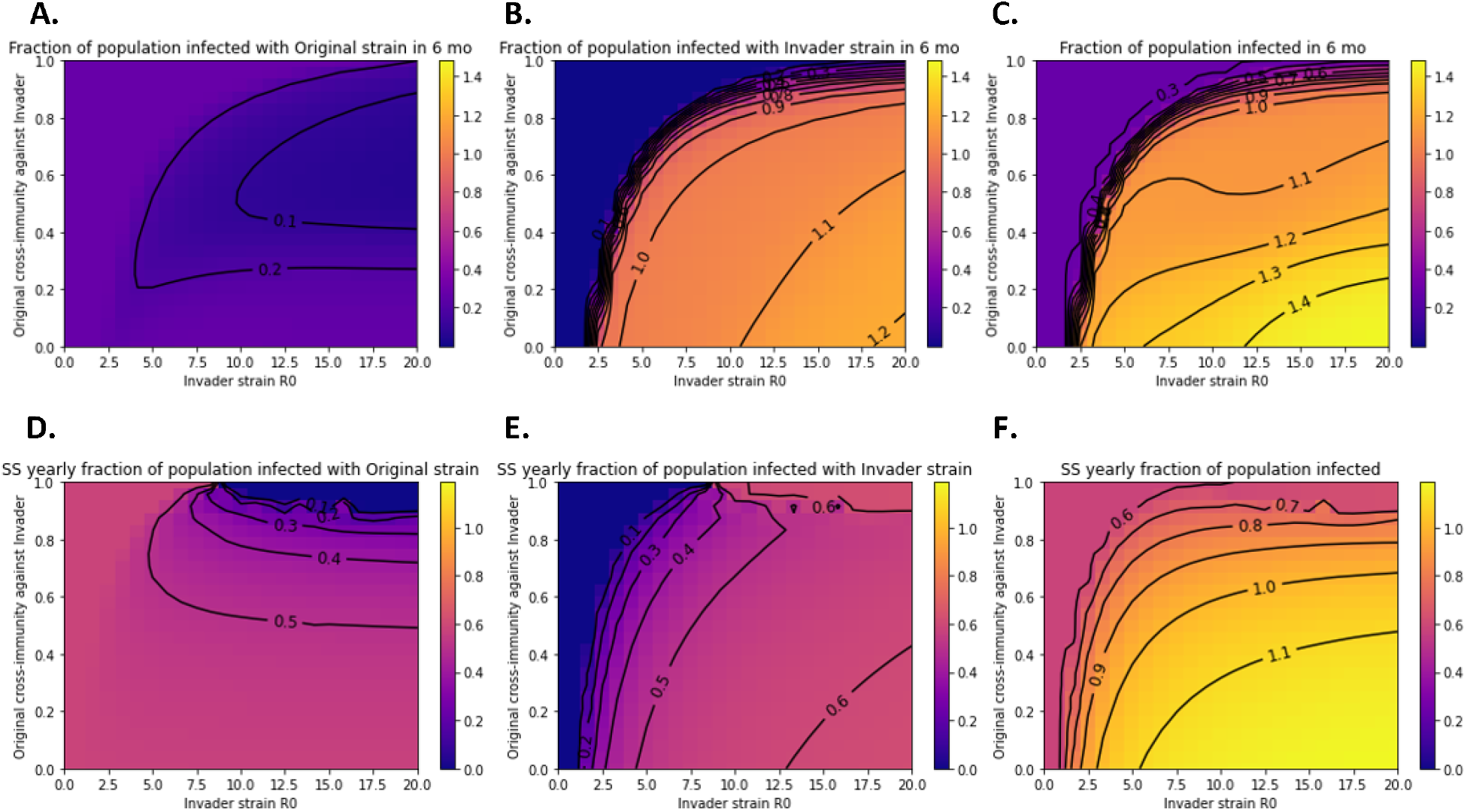
Short-term and long-term transmission trends after introduction of invader strains with varying transmissibility and symmetric cross-immunity properties. The total number of infections is expressed as a fraction of the population size. Fractions greater than 1 indicate reinfection. Total infections over six months due to **A)** the original strain, **B)** the invader strain, or **C)** both summed together. Total infections over one year at steady-state for **D)** the original strain, **E)** the invader strain, or **F)** both summed together. Colormaps are matched across subpanels A-C and D-F.

### Unilateral immune evasion favors succession

Figure 4 explores invasion scenarios in which the invader strain evades the original strain’s immunity but exerts full cross-immunity against the original strain. While Figure 2 demonstrates that succession is rare if immune evasion is perfectly symmetric, Figure 4 indicates that unilateral immune evasion favors succession, especially if the R_0_ of the invader strain is equal to or greater than the original strain’s R_0_. However, the conditions under which invasion succeeds – represented by both co-circulation and original strain extinction outcomes – are identical to the mutual symmetric immune evasion scenario.

**Figure 4.**
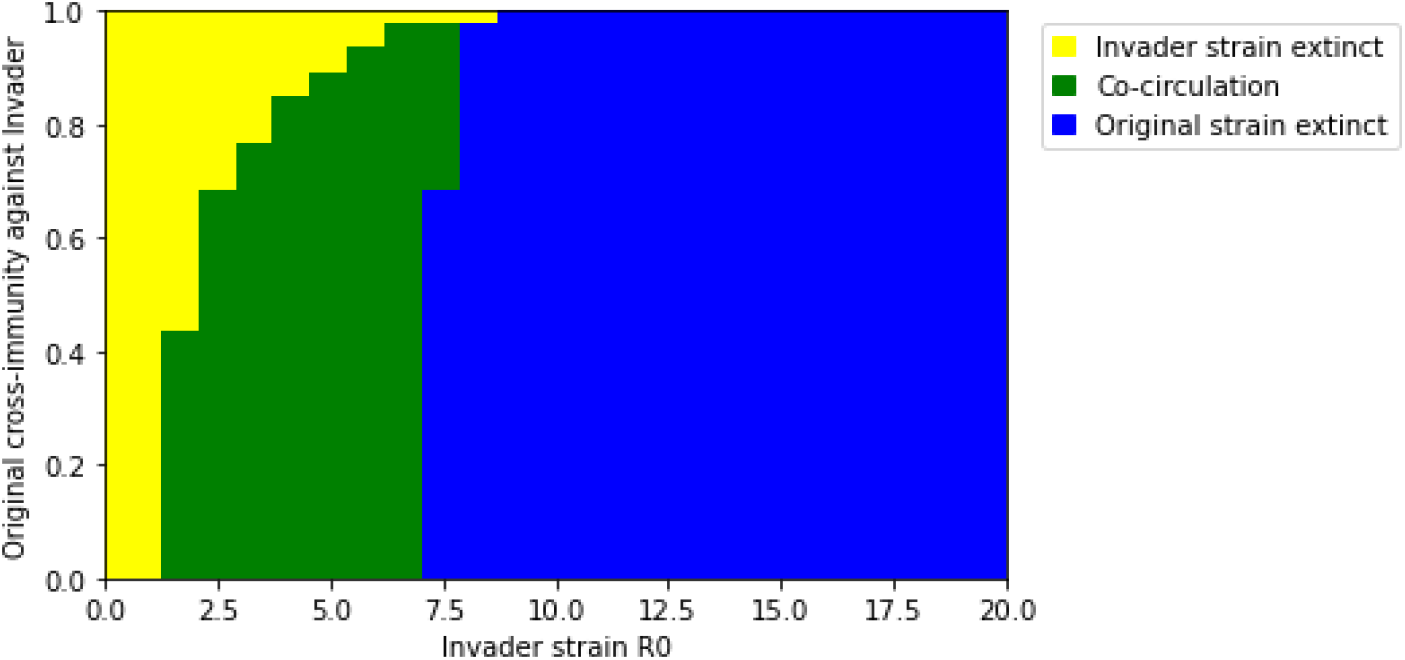
Invasion outcomes for novel variants with varying transmissibility and degrees of asymmetrical immune evasion.

In the short-term, transmission dynamics after emergence of an invading strain in the asymmetric scenario are similar to the symmetric scenario (Figures 5A-C). This is intuitive: the difference between the two scenarios is in the nature of the invading strain’s immunity, which is not yet present in the short-term. In the long term, immunity created by the invading strain constrains spread of the original strain, in many cases leading to its extinction (Figures 5D-F). This reduces transmission of the pre-existing strain with limited benefit to the invading strain, except in cases where the invading strain’s dominance is weaker. As a result, long-term, overall transmission is not substantially impacted by successful invasion when cross-immunity induced by the invading strain against the pre-existing strain is strong.

**Figure 5.**
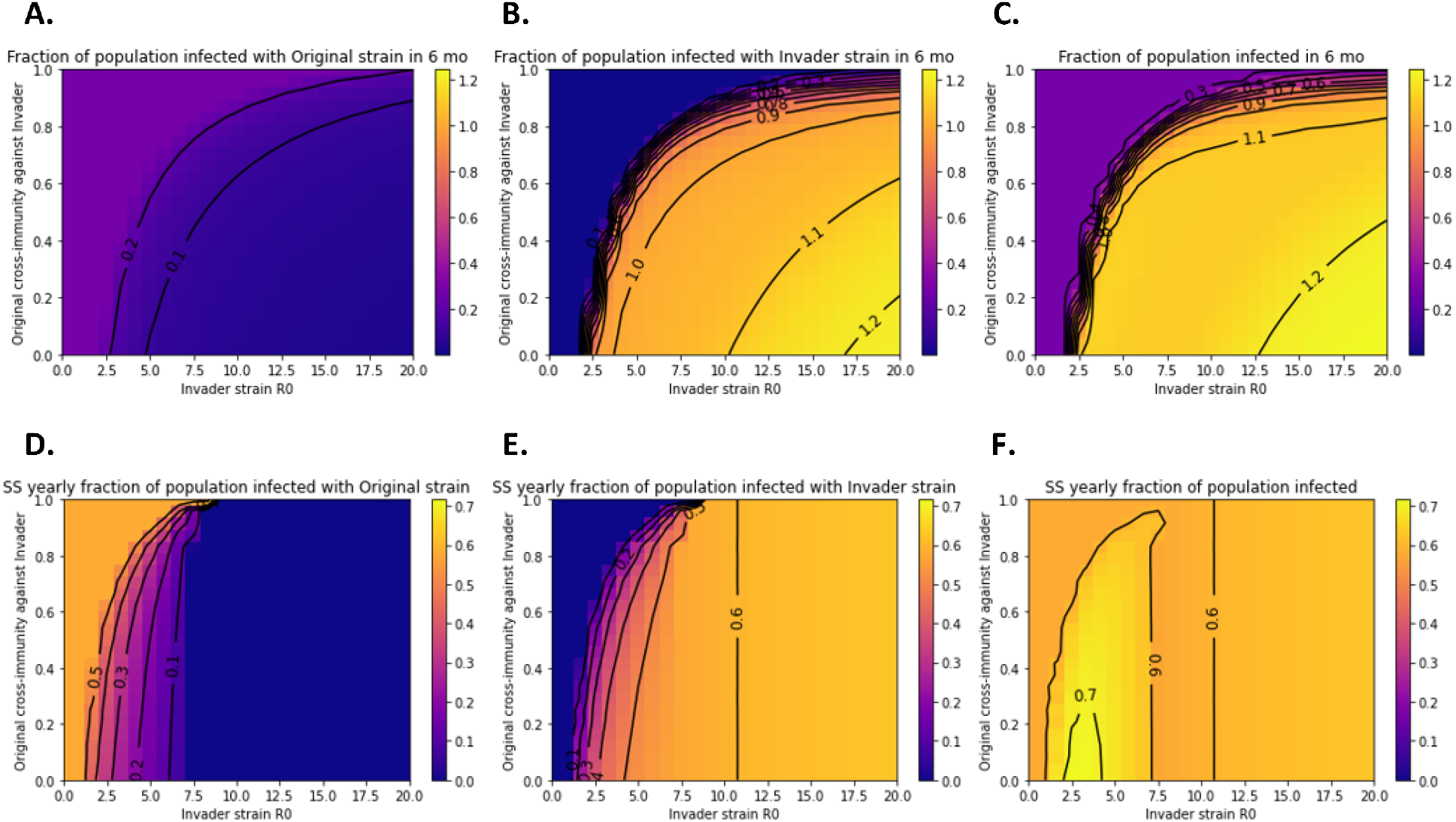
Short-term and long-term transmission trends after introduction of invader strains with varying transmissibility and unilateral cross-immunity properties. The total number of infections is expressed as a fraction of the population size. Fractions greater than 1 indicate reinfection. Total infections over six months due to **A)** the original strain, **B)** the invader strain, or **C)** both summed together. Total infections over one year at steady-state for **D)** the original strain, **E)** the invader strain, or **F)** both summed together. Colormaps are matched across subpanels A-C and D-F.

### Duration of invader strain’s immunity does not impact its potential to invade

We also simulated invasion attempts by strains that exert shorter-term immunity. We found that in the absence of antibody evasion, the duration of immunity induced by the invading strain does not impact its ability to establish (Figure 6). This is consistent with the intuition that immunity induced by the invader strain does not yet exist at the time of invasion and likewise does not impact invasion success. Additionally, a strain that induces less durable immunity achieves no selective advantage in the absence of antigenic drift because the shorter immunity increases the reinfection potential of both strains equally. Additionally, we note that variation in this property in the absence of immune evasion does not support coexistence of strains. However, strains with shorter duration of immunity may transmit at much higher levels in the long term (Figure S1).

**Figure 6.**
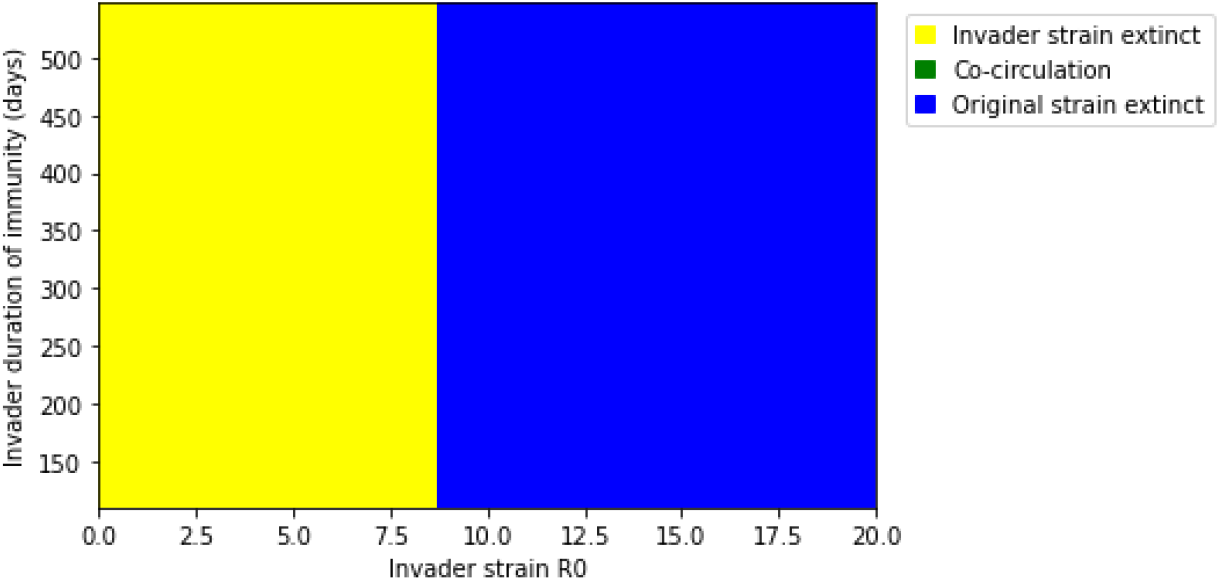
Outcomes for invasion scenarios in which the novel strain induces less durable immunity than the original strain. The duration of immunity against wild-type SARS-CoV-2 infection is estimated to be 550 days [42].

## Discussion

Based on the epidemiological modeling analyses in this work, we determine that cross-immunity between circulating and emergent strains is expected to be a major driver of the ability of invading strains to successfully establish as well as short-term and long-term transmission trends. This includes the possibility of stable co-circulation of antigenically distinct strains (serotypes). Under conditions of widespread anti-SARS-CoV-2 immunity, antibody evasion likely provides an evolutionary advantage. Increases in immune evasiveness can be expected to drive selection and transmission to a greater extent than further increases in intrinsic transmissibility (R_0_) alone.

Pre-existing immunity against emergent strains – by definition, imposed by the original strain – and intrinsic transmissibility shape the invasion potential and short-term transmission of new strains. This is the cross-immunity of the original strain against the invader strain. Strains with antigenic drift that weakens pre-existing immunity are likely to be strongly selected for and drive significant increases in short-term transmission. In this way, ongoing viral evolution is likely to contribute to long-term high and volatile levels of SARS-CoV-2 transmission. Notably, even poorly transmissible (low R_0_) strains may successfully invade if they sufficiently evade pre-existing antibodies.

Although evasion of pre-existing immunity dictates which strains will successfully emerge, the properties of immunity induced by successful invading strains will determine their long-term impact on overall transmission levels. For example, we have shown that invader strains inducing less durable immunity sustain greater long-term transmission if invasion is successful.

Additionally, weak cross-immunity of the invader strain against the original strain can permit co-circulation of the two strains (“serotypes”). This may significantly increase long-term transmission levels. The existence of serotypes for other human pathogens (e.g. dengue) has made their control more complicated, as tests and vaccines have to be created to match each serotype [46]. Co-circulation of SARS-CoV-2 serotypes would increase demand on already-strained global sequencing systems for tracking viral evolution and may exacerbate the challenge of matching vaccines and therapeutics to circulating variants. Serotypes are also thought to increase the risk of antibody-dependent enhancement (ADE), as has been observed for dengue [47]. Although ADE is not thought to be a risk for SARS-CoV-2 at present, the cocirculation of two or more SARS-CoV-2 serotypes raises the risk of increased virulence of COVID-19 through this mechanism [48].

These findings highlight the importance of measuring cross-immunities between pre-existing and emergent variants to forecast the likelihood of invasion success and short-term and long-term implications of a successful invasion. Although real-world cross-immunity is challenging to measure, neutralizing potency of post-immunization sera has proven to be a practical correlate [49]. Many studies assess the neutralization potency induced by vaccines against emerging variants [50,51], while some studies have also assessed the neutralization potency raised by circulating strains against emerging variants [52–54]. Using models linking neutralizing antibody potency to extent of protection, this data can be leveraged to estimate cross-immunity against invading variants [55]. This is important for predicting which variants are likely to establish successfully and significantly increase short-term transmission.

However, data is often lacking regarding the neutralization potency induced by emergent variants against pre-existing variants. Measuring the neutralizing potency of invading variant infectee sera against pre-existing variants could reveal the risk of long-term co-circulation. In this work, we have demonstrated that co-circulation may follow apparent declines in the transmission of the original strain during high post-emergence transmission of the invader strain. This underlines the importance of in vitro or clinical data regarding cross-immunity of invading strains against pre-existing strains. The likelihood of co-circulation may not be obvious in early epidemiological data, at which point interventions are most likely to succeed.

There are a few limitations to this analysis. Firstly, we did not simulate the impact of vaccines in this analysis. In our prior work [56], we have explored the impact of vaccines on SARS-CoV-2 evolutionary dynamics; we determined that the impact of vaccines on evolutionary dynamics is limited due to minimal vaccinal protection against infection. Secondly, our SIRS model implementation categorizes individuals into four immunological groups: fully susceptible, immune to original strain and susceptible to invader strain, immune to invader strain and susceptible to original strain, and immune to both strains. As we have explored in other analyses using agent-based models, the immunity landscape is more complex than this SIRS model can capture. This complexity arises due to heterogeneity in individual exposure to infection and vaccination, interindividual variability in antibody durability, and neutralizing antibody build-up over successive infections and vaccinations [42,57]. In particular, we surmise that the build-up of neutralizing potency over successive infections may cause the apparent cross-immunities between strains to not be fixed over time. This may result in additional competitive dynamics in the long term. Lastly, our analysis addresses competition between two strains for hosts in short and long timeframes. In reality, multiple strains may be in competition simultaneously, and the emergence of further invader strains may occur before the “long-term” two-strain dynamics explored here are fully realized [58,59]. We also acknowledge the possibility of short-term “pseudo” co-circulation, in which strains may appear to co-circulate in the short-term before sufficient infections have occurred to impose immunological constraints. Nevertheless, our work provides a basis for understanding basic competition dynamics between any two strains.

With these limitations in mind, the purpose of this analysis is to broadly identify variant properties conducive to invasion and to highlight the risk and implications of co-circulation of SARS-CoV-2 strains.

Our work demonstrates that viral evolution is a key driver of transmission rates at this stage of the pandemic, as invasion events are likely to drive extensive transmission in the short-term. We emphasize the value of early data characterizing the antibody evasion of and immune responses to emergent variants to inform SARS-CoV-2 vaccination and other mitigation strategies. Our work also suggests the emergence of SARS-CoV-2 serotypes as a further potential risk of the current public health strategy. In sum, the results presented here point to the likelihood of high baseline SARS-CoV-2 transmission, with the possibility of transient increases in transmission (“waves”) driven by successive (or co-circulating) waves of immune-evasive variants. We identify inter-strain cross-immunity as an important variable for predicting outcomes and gauging risk in this seemingly unpredictable scenario.

## Data Availability

All data produced in the present study are contained in the manuscript. Code used to produce the results is available on Github.

https://github.com/madistod/two-strain-SEIR

## Supplement

**Figure S1.**
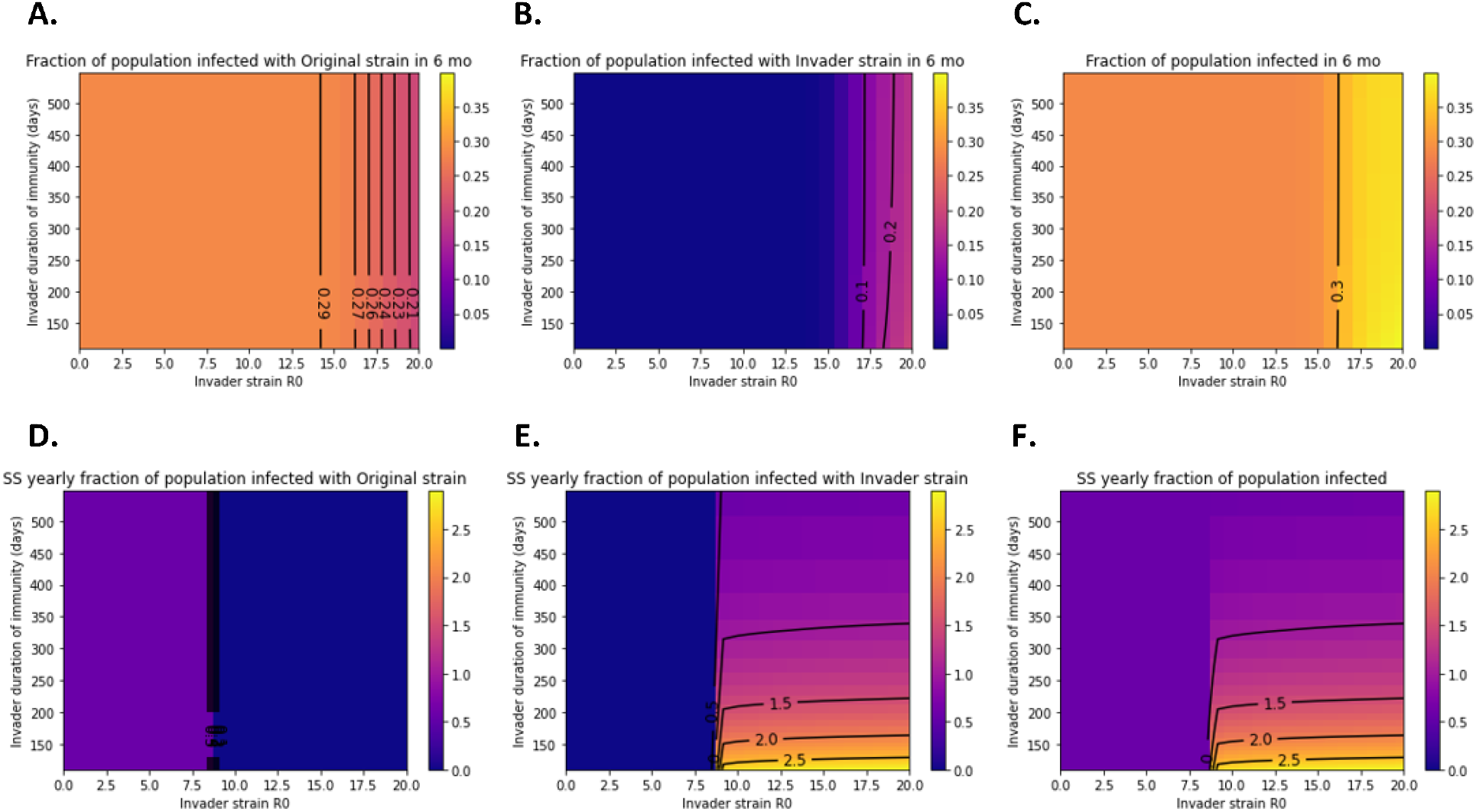
Short-term and long-term transmission trends after introduction of invader strains with varying transmissibility and durability of immunity. The total number of infections is expressed as a fraction of the population size. Fractions greater than 1 indicate reinfection. Total infections over six months due to **A)** the original strain, **B)** the invader strain, or **C)** both summed together. Total infections over one year at steady-state for **D)** the original strain, **E)** the invader strain, or **F)** both summed together. Colormaps are matched across subpanels A-C and D-F.

